# How many microbial species are there in human tumor and normal tissues?

**DOI:** 10.1101/2024.04.16.24305922

**Authors:** Zhanshan (Sam) Ma

## Abstract

The study of tissue microbiomes is a recent endeavor in human microbiome research, particularly in the area of blood microbiomes. This is primarily due to their low biomass, which presents inadvertent operational contamination as a significant experimental obstacle. The critical role of the tissue microbiomes in cancer development has brought this topic to the forefront of cancer research. However, a fundamental question regarding the potential biodiversity, as stated in the title, has not been addressed to our knowledge. In this study, we estimate the potential microbial diversity or “dark” biodiversity in human tumor and normal tissues using the Diversify-Area Relationship (DAR) method (Ma 2018, 2019) based on large datasets from TCGA (The Cancer Genome Atlas) database (Poore *et al*. 2021). We found that the total species richness (number), typical species equivalents (number), and dominant species equivalents (number) of tumor tissues are approximately 1948, 36, and 22, respectively. Among the total species richness, the proportions of archaea, bacteria, and viruses are about 3%-5%, 78%-79%, and 17%-18%, respectively. Moreover, the tissue species richness is approximately 12.5% of skin microbiomes, and 25% of gut microbiomes. We also found that tumor growth does not significantly influence the global or pan-tumor scale diversity, which means that the previous numbers also represent the potential microbial diversity of human tissues, including blood. On a local or single cancer-type scale, tumors may influence the potential diversity in approximately 5% of cases. We hypothesize that, globally, local diversity variations would offset each other.

**Lay Summary:** This study focused on microbiomes - the tiny microbes that live in our tissues, especially blood. Studying them is challenging due to their low biomass and risk of contamination. Microbiomes may play a key role in cancer, but their diversity within tissues remains unclear. Using diversity-area relationship modeling with data from “The Cancer Genome Atlas” database, we estimated the potential microbial diversity of human tumor and normal tissues. We found approximately 1,948 microbial species in tumors, comprising archaea (5%), bacteria (78%) and viruses (17%). However, dominant or common microbial species number only about two to three dozen. Tissue microbe diversity was 12.5% of that found in skin microbiomes and 25% of gut microbiomes. Tumor growth did not significantly impact overall diversity. Therefore, the previous diversity numbers also represent the microbiome diversity of general human tissues and blood. However, some cancer types may affect it locally. While local diversity changes can occur, globally these variations between tissues likely balance out.

## Introduction

Recent reviews have proposed that polymorphic microbial communities, also known as heterogeneous microbiomes, could represent an emerging cancer hallmark or enabling characteristic. Cancer hallmarks refer specifically to core biological capabilities that drive tumor development and progression. In contrast, enabling factors facilitate the acquisition of hallmark abilities without being hallmarks themselves. In his 2022 synthesis review, Hanahan (2022) suggested phenotypic flexibility and disrupted development as potential additional hallmark capabilities, adding to the currently well recognized eight hallmarks that he co-proposed about a decade ago. He also posited that non-mutational epigenetic changes and polymorphic microbiomes act as distinctive enabling characteristics that help tumors develop hallmark properties. Similarly, Lythgoe et al (2022) directly referred to microbes as an emerging hallmark of cancer in their 2022 review. Hanahan’s distinction of microbiomes as enabling factors rather than hallmarks reflects the slightly different level of the recognition of the critical role of microbes in cancer research. Regardless of the ultimate position on the role of microbiomes in cancer development and progression, their critical importance has been well recognized in the recent decade. For example, microbiome-immune cell interactions likely influence a tumor’s ability to evade immune destruction, one of the currently established key cancer hallmarks. Heterogeneous microbiomes can profoundly impact cancer phenotypes by differentially affecting hallmark processes. Obviously, the microbiome also contributes heterogeneity between patients (Sepich-Poore *et al*. 2021).

Sepich-Poore et al. (2021) discussed the role of microbes in cancer from a historical and modern perspective. Early studies dating back 4000 years linked cancer to microbes (Sepich-Poore et al. 2021). One of the first clinical studies in 1868 observed tumor regressions in streptococcus-infected patients, providing an early demonstration of immunotherapy. However, these claims faced reproducibility issues and toxicity concerns over the next century. While the viral theory of Rous sarcoma virus gained traction in 1911, decades of searching failed to find viruses causing human cancers. Instead, somatic mutations are now primarily linked to many cancers. Recent studies reconsider the importance of bacteria and fungi in cancer and immunotherapy through immune-mediated mechanisms (Sepich-Poore et al. 2021). The human microbiome project enabled large-scale metagenomic sequencing, fueling renewed interest in microbe-cancer relationships over the past decade. Landmark studies in 2020 (Nejman *et al*. 2020, Poore et al. 2020) provided experimental and computational evidence, signifying a breakthrough in understanding the cancer microbiome’s role and potential for improving immunotherapy.

Of the estimated 10^12^ microbial species on Earth, only 11 have been identified as human carcinogens (Sepich-Poore et al. 2021). Although only a small number of microbes directly cause cancer, many appear to promote tumor development and progression through immune-mediated interactions, a process known as the immuno-oncology-microbiome axis. Key questions in this field include the roles of microbes - whether they are causal, complicit, or merely passive bystanders, and the current understanding of intertumoral microbes (Sepich-Poore et al. 2021). This article seeks to tackle one outstanding issue, part of the open problem regarding intertumoral or tissue microbiomes, seemingly straightforward as indicated by the title: the diversity of microbial species found in human tumor and normal tissues. Our primary focus is on the overall diversity of microbes in human tissues, a topic that, to our knowledge, has not been previously explored. While comprehensive studies have been conducted on local microbial diversity, known as alpha-diversity, and even beta-diversity, which evaluates regional or inter-locality diversity (including our own studies, Ma 2024a, b), the global diversity of microbes in human tissues remains an uncharted territory.

The challenge of estimating total or global biodiversity was first posed in the 19th century (Watson 1835), during the era of Charles Darwin when naturalists were passionately cataloguing the flora and fauna of our planet. A straightforward approach might be to aggregate the data collected by naturalists worldwide. However, two significant issues arise with this approach. Firstly, the vastness of the Earth makes it impossible for naturalists to reach every location where organisms exist. Secondly, there may be overlapping data in the catalogues compiled by different naturalists, whether from the same or different regions. A potential solution to the first issue could be implementing effective sampling schemes, as it is neither feasible nor necessary to conduct exhaustive counts of species numbers, especially considering the ongoing processes of speciation and extinction. The second issue could be addressed through an automated algorithm designed to eliminate overlaps. The complexity of these issues necessitates the use of statistical or stochastic algorithms to handle the challenges of sampling, stochasticity, and overlap (e.g., Connor and McCoy 1979, Chao et al. 2014).

The first attempt to estimate biodiversity was made by British plant biogeographer and evolutionist, Hewett Cottrell Watson (1804–1881), who introduced the concept known as the species-area relationship (SAR) (Watson 1835). The SAR correlates the accumulation of species numbers (S) with the size of the area (A) where the species are found, using a power function (*S* = c*A*^*z*^). Intuitively, a larger area should be inhabited by more species, or in other words, the larger the area a naturalist surveys or samples, the more species they should collect. For this reason, SAR is also referred to as the collector’s (or naturalist’s) accumulation curve. The SAR can be transformed into a linear relationship on a logarithmic scale, i.e., ln(*S*) = ln(*c*) + z*ln*(*A*), which makes the increasing correlation between species number and area size even more apparent. Since Watson’s pioneering work, the SAR has been extensively and intensively studied in community ecology and biogeography (e.g., Preston 1962, Connor and McCoy 1979, Rosenzweig 1995, Plotkin et al. 2000, Ulrich & Buszko 2003, Tjørve & Tjørve 2008, Triantis et al. 2012). For instance, the SAR served as a foundational model for MacArthur and Wilson’s (1967) island biogeography theory, which significantly influenced community ecology in the 1960s and 1970s. In practical terms, the SAR has arguably become the most critical model in conservation biology, particularly for protecting biodiversity and endangered species, influencing decisions such as the appropriate size of conservation zones for endangered species (Rosenzweig 1995).

Despite its widespread applications, the classic SAR model presents two issues. The first pertains to the ever-increasing or decreasing nature of the power function in the SAR model, which lacks “saturation” points or extreme (maximum or minimum) values. Given the Earth’s finite size, it’s reasonable to argue that the number of species should also be finite. To address this issue, Plotkin et al. (2000) and Ulrich & Buszko (2003) introduced the power law model with exponential cutoff (PLEC) and power law with inverse exponential cutoff (PLIEC), both of which incorporate saturation points (extreme values). With PLEC or PLIEC, the SAR curve can reach saturation or maximums, corresponding to the maximum number of species on Earth, or alternatively, to the maximum number of species in a specific region, such as human tissues.

The second issue associated with the classic SAR relates to the definition of biodiversity, which, in its simplest form, is the number of species in a region (area), known as species richness (S or R). The problem with this simplified definition is that it fails to account for the fact that not all species are equal: some are abundant (such as ants and many insects), while others are not only rare (e.g., pandas and tigers), but also potentially more valuable. In the context of this study, as previously mentioned, only 11 microbial species have been identified as human carcinogens or oncomicrobes according to the International Association for Cancer Registries (IACR) (cited in Sepich-Poore et al. 2021). It is clear that different microbial species in human tissues have varying oncological significances, and our interest should lie in estimating the numbers of microbial species at various levels of significance.

Indeed, there are numerous diversity metrics (indexes) beyond the simplest measure of species richness. So many, in fact, that practitioners often find themselves overwhelmed by the multitude of choices, lacking a solid standard to guide their proper applications (Magurran 2013, Henderson 2021). Among these many diversity metrics, Shannon’s entropy and Simpson’s index are two of the most commonly used. It might seem that a simple substitution of species richness in the classic SAR with these metrics would solve the problem, but the solution is not that straightforward. Diversity metrics such as Shannon entropy and Simpson’s index are not only incompatible with each other, but they also do not scale in terms of simple mathematical functions like power-law models. This complexity may explain why a breakthrough in this area has been elusive for a long time.

One diversity metric that possesses such appropriate properties is the so-called Hill numbers, first introduced as biodiversity metrics by Hill (1973) from economics. However, it did not garner the attention it deserved among ecologists until its rediscovery by Chao et al. (2014), possibly due to the somewhat abstruse interpretations Hill (1973) used to explain its central concept of “numbers equivalent of elements” in economics. The accomplishment of Hill numbers is actually similar to linking the US dollar to gold at the rate of $35 per ounce under the Bretton Woods system, as per Ma & Li (2024). Hill numbers are now considered the most suitable system for biodiversity metrics, unifying Shannon, Simpson, and other diversity indexes. Against this backdrop, Ma (2018a, 2019) extended the classic SAR into the Diversity-Area Relationship (DAR) using Hill numbers, and also to the Diversity-Time Relationship and Diversity-Time-Area Relationship (DTAR). The extensions also incorporated the adoption of PLEC and PLIEC as DAR models and derivations of maximal accrual diversity (MAD). The MAD or D_max_ essentially represents the potential diversity or ‘dark’ diversity, accounting for species that may be locally absent but exist in the regional species pool (and may therefore immigrate at a certain time). In the context of this study, it means that MAD can account for microbial species that are absent in some individuals while present in others.

With the DAR-PL and DAR-PLEC models, and leveraging a recent breakthrough in computational approaches to distill large datasets of tumor tissue microbiomes, we can address the question raised in the article title. This has been made possible thanks to the revolutionary AI-machine learning approach by Poore et al. (2020), who produced a substantial dataset of tissue microbiomes from the TCGA (The Cancer Genome Atlas) database. To enhance the robustness of our estimations, we employ extensive permutation tests to manage the inherent stochasticity (uncertainty). Figure 1 and Table 1 provide a roadmap of our study.

**Table 1.**
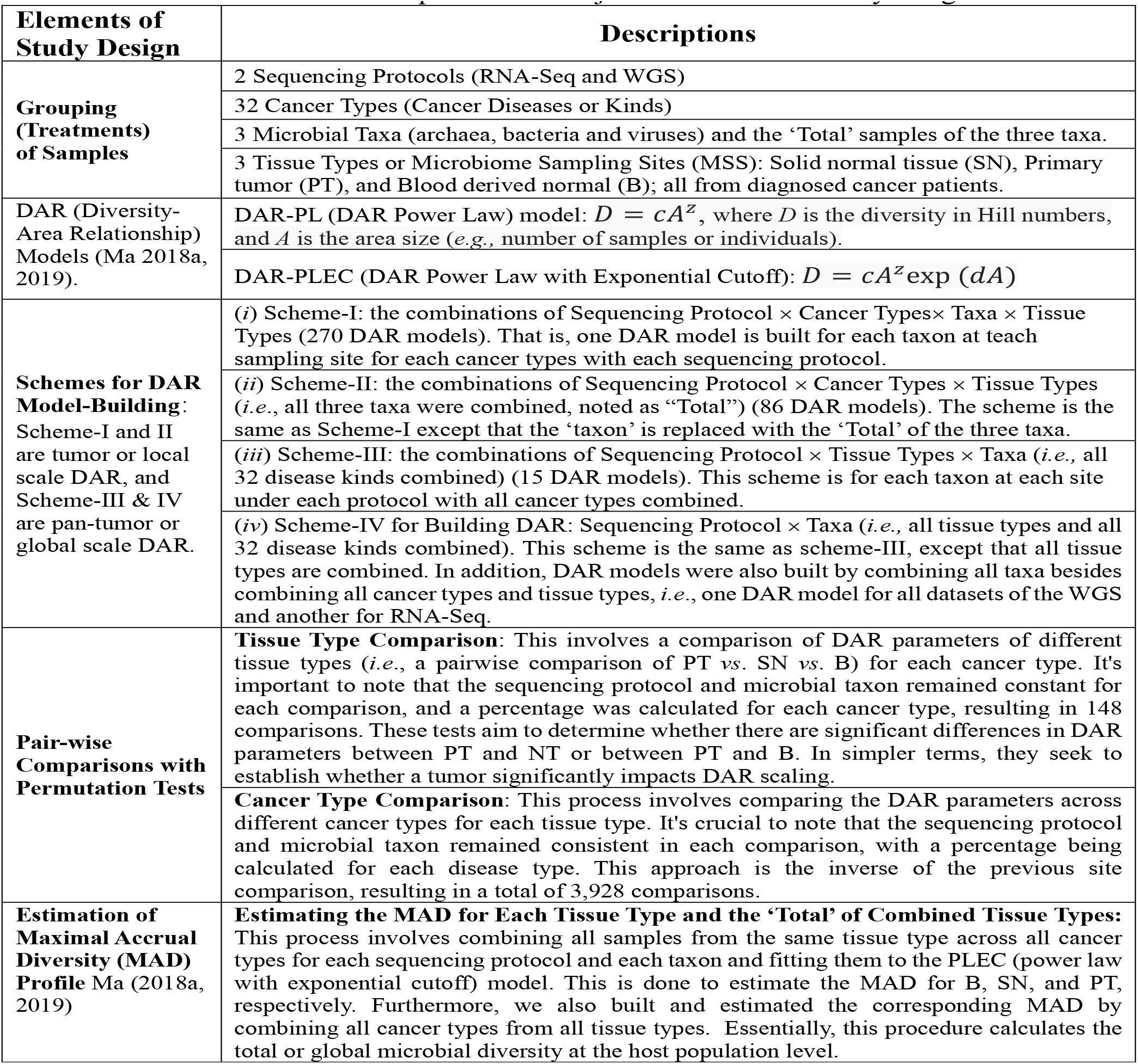
Brief description of the major elements of the study design.

**Fig 1.**
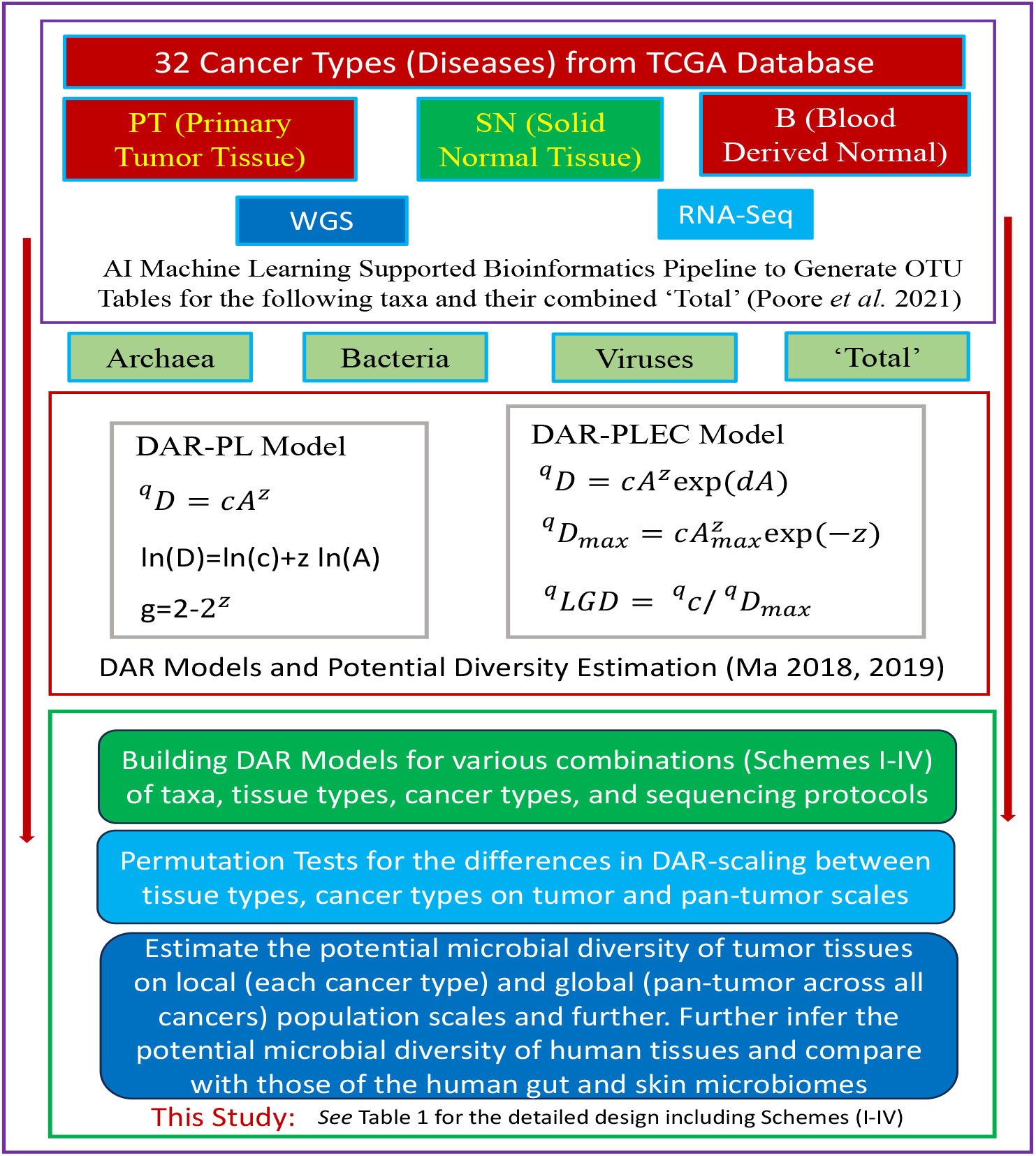
A diagram for illustrating the study design and its relationships with previous foundations works (also *see* Table 1 for the supplementary interpretations on the design). Legends for the equations: ^*q*^*D* is the diversity in Hill numbers at diversity order *q*=0, 1, 2, 3; *A* is the ‘area’ size (*e*.*g*., the number of samples or individuals); ^*q*^*D*_max_ is the MAD (maximal accrual diversity) of diversity order *q*, the so-termed potential or ‘dark’ diversity, which includes both local species and species that are absent locally but present in regional species pool.

## Material and Methods

### The Microbiome Dataset of Tumor Tissues and Study Design

The cancer microbiome datasets encompass 32 different types of cancer, each represented by microbial samples taken from one to three sample sites: primary tumor (PT), solid normal tissue (SN), and blood-derived normal (B). Each disease’s tissue site is considered a specific group or treatment, with samples collected from a cohort of patients diagnosed with the same cancer type. To avoid issues related to small sample sizes, groups with fewer than 15 samples were excluded from the analysis. This resulted in a dataset of 17,066 samples across 32 cancer types for further host-population level diversity analysis. It’s crucial to note that all microbiome samples were obtained from patients with confirmed cancer diagnoses. Figure 1 sketches out the study design, and Table 1 provides supplementary interpretations of the design.

## Results

### Analysis of Tumor Microbiome Diversity Scaling

We initially constructed tumor microbiome diversity scaling models, specifically DAR-PL (diversity-area relationship with power law) and DAR-PLEC (DAR with power law with exponential cutoff), based on Scheme-I and II as outlined in Table 1 and Figure 1, with the results documented in Tables S1 and S2. The tumor tissue microbiome datasets were well-fitted by all PL and PLEC models, as indicated by a *P*-value less than 0.05 (refer to Table S1 for RNA-Seq and Table S2 for WGS). Besides the *P*-values, Tables S1 and S2 also present all DAR parameters.

### Comparing tissue types in their diversity scaling parameters within the same cancer type

While Tables S1 and S2 list the DAR parameters for the RNA-Seq and WGS datasets respectively, Table S3 exhibits the results of permutation tests comparing different tissue types of the same cancer type. Specifically, it shows the *P*-values from tests conducted for each DAR parameter.

Except for the D_max_, A_max_, and LGD parameters, the differences between tumor sites in other DAR scaling parameters are negligible (only a few comparisons showed statistically significant differences). Even for Dmax, Amax, and LGD, the percentage of statistically significant differences generally falls below 5% (see Table S3 and Table S4). An exception was observed in Kidney Renal Clear Cell Carcinoma, but the differences were only around 10% in most cases. These findings suggest a universal diversity scaling across different microbiome sampling sites (tissue types). In other words, it appears that, within the same tumor type, diversity scaling parameters are not influenced by tumor site. This is in strong contrast with the findings from alpha-diversity and beta-diversity, as elaborated in the discussion section.

### Comparing cancer types in their diversity scaling parameters for the same tissue type

Different from the previous comparisons of different tissue types (microbiome sampling sites, e.g., primary tumor [PT] vs. solid normal [SN]) within the same cancer type (e.g., lung cancer) in Tables S3 and S4, Tables S5 and S6 exhibit the results from comparing different cancer types (e.g., lung cancer vs. breast cancer) for the same tissue type (*e*.*g*., PT).

First, compared to the previous tests of tissue types, statistically significant differences among cancer types are more prevalent in terms of the RNA-Seq protocol, but less prevalent or similar to the previous comparisons of tissue types for the WGS protocol. In other words, the sequencing protocols appear to make a significant difference in this case.

With the RNA-seq protocol, statistically significant differences among cancer types were particularly prevalent for D_max_ (mostly approximately 16-34% on average). For the other scaling parameters (except D_max_), statistically significant differences were more prevalent only under diversity order *q*=0 or species richness (mostly approximately 10-34% on average). For other diversity orders, statistically significant differences in the other scaling parameters (mostly below 5% on average) were more prevalent than in the previous comparisons of tissue types, but less prevalent than in the comparisons of D_max_.

Also, with the RNA-Seq protocol, statistically significant differences among cancer types appear to be more prevalent for primary tumor (PT) than for solid normal (SN), approximately one-third more prevalent.

With the WGS protocol, statistically significant differences among cancer types seem to be more prevalent only in terms of Dmax (mostly approximately 16-40% on average) compared to the previous comparisons of cancer tissue types (mostly under 10%). In terms of the other scaling parameters, the differences are similar to the previous cases (mostly under 5%).

### Pan-tumor Microbiome Diversity-Scaling Analysis

Given the nearly universal invariance in major DAR scaling parameters, especially *z*, it is justified to combine all cancer disease types, and even tissue types to analyze diversity scaling across cancer/tissue types. Table S7 exhibits the results of fitting the DAR-PL and DAR-PLEC models with all cancer types and/or tissue types combined, based on the designs of Scheme-III and IV in Table 1. Table 2 below excepts the key DAR parameters for convenience of illustration, with the combined tissue types and cancer types. We further compared the DAR parameters with permutation tests based on 1,000 repetitions of re-sampling, and the test results were exhibited in Table S8. It turned out that no statistically significant differences were detected in any of the comparisons (P<0.05) between different tissue types in Table S8. That is, there are no statistically significant differences between tumor tissue types (PT, SN, or B) in any of the DAR parameters for the models built based on Scheme-III and IV. In other words, on a pan-tumor basis, diversity scaling makes no difference between tissue types. This suggests that tumors do not significantly influence the total microbiomes on a pan-tumor basis or across cancer types.

**Table 2.** The DAR modeling parameters from the combined datasets of all tissue types and cancer types, for each of the three taxa (Archaea, Bacteria, and Viruses) under each sequencing protocol (RNA-Seq or WGS), excerpted from Table S7.

| Sequencing Protocol | Taxon | Diversity Order | Power Law (PL) |  | PL with Exponential Cutoff (PLEC) |  |  |  |  |  |
| --- | --- | --- | --- | --- | --- | --- | --- | --- | --- | --- |
|  |  |  | z | ln(c) | z | d | ln(c) | A<sub>max</sub> | D<sub>max</sub> | LGD (%) |
| RNA-seq | Archaea | $q = 0$ | 0.011 | 4.53 | 0.031 | 0 | 4.398 | 4920 | 102.9 | 90.2 |
| | | $q = 1$ | 0.001 | 1.857 | 0.003 | 0 | 1.844 | 5942 | 6.5 | 99.3 |
| | | $q = 2$ | -0.002 | 1.265 | -0.006 | 0 | 1.292 | 6173 | 3.5 | 103.2 |
| | | $q = 3$ | -0.003 | 1.055 | -0.01 | 0 | 1.096 | 6778 | 2.8 | 104.4 |
| | Bacteria | $q = 0$ | 0.003 | 7.304 | 0.009 | 0 | 7.264 | 4729 | 1528.3 | 97.2 |
| | | $q = 1$ | 0.029 | 3.335 | 0.063 | 0 | 3.113 | 7399 | 36.6 | 77.2 |
| | | $q = 2$ | 0.041 | 2.699 | 0.092 | 0 | 2.375 | 6748 | 21.8 | 69.8 |
| | | $q = 3$ | 0.043 | 2.462 | 0.099 | 0 | 2.104 | 6486 | 17.5 | 68.7 |
| | Viruses | $q = 0$ | 0.138 | 4.472 | 0.16 | 0 | 4.331 | 29269 | 329.6 | 26.7 |
| | | $q = 1$ | 0.026 | 2.731 | 0.056 | 0 | 2.539 | 8821 | 19.6 | 88.9 |
| | | $q = 2$ | 0.038 | 1.857 | 0.07 | 0 | 1.663 | 13539 | 9.2 | 88.7 |
| | | $q = 3$ | 0.038 | 1.562 | 0.066 | 0 | 1.388 | 10355 | 6.8 | 87.9 |
| WGS | Archaea | $q = 0$ | 0.014 | 4.521 | 0.037 | 0 | 4.397 | 1730 | 102.9 | 89.4 |
| | | $q = 1$ | -0.001 | 1.015 | -0.004 | 0 | 1.028 | 2962 | 2.7 | 101.4 |
| | | $q = 2$ | -0.002 | 0.546 | -0.006 | 0 | 0.567 | 2449 | 1.7 | 102.1 |
| | | $q = 3$ | -0.002 | 0.433 | -0.005 | 0 | 0.452 | 2707 | 1.5 | 102 |
| | Bacteria | $q = 0$ | 0.007 | 7.273 | 0.021 | 0 | 7.199 | 1684 | 1532.6 | 94 |
| | | $q = 1$ | 0.013 | 2.913 | 0.028 | 0 | 2.837 | 4431 | 20.6 | 90.1 |
| | | $q = 2$ | 0.004 | 2.128 | 0.016 | 0 | 2.059 | 2766 | 8.7 | 95.8 |
| | | $q = 3$ | 0.002 | 1.887 | 0.011 | 0 | 1.837 | 6325 | 6.7 | 98.3 |
| | Viruses | $q = 0$ | 0.136 | 4.669 | 0.15 | 0 | 4.595 | 99977 | 367.6 | 29.6 |
| | | $q = 1$ | 0.034 | 2.982 | 0.043 | 0 | 2.94 | 2991 | 25.8 | 81.4 |
| | | $q = 2$ | 0.049 | 2.028 | 0.065 | 0 | 1.952 | 3418 | 11.2 | 73.4 |
| | | $q = 3$ | 0.046 | 1.67 | 0.069 | 0 | 1.559 | 11039 | 7.8 | 73.3 |
| Range across RNA-WGS protocols | Archaea | $q = 0$ | 0.04±0.014 | 4.375±0.08<sub>1</sub> | 0.08±0.023 | 0±0 | 4.207±0.1 | 1921±751 | 102.2±0.5 | 79.2±5.4 |
| | | $q = 1$ | 0.002±0.003 | 1.354±0.17<sub>8</sub> | 0.003±0.00<sub>7</sub> | 0±0 | 1.351±0.17 | 3673±1547 | 4.4±0.9 | 99.5±1.8 |
| | | $q = 2$ | 0±0.003 | 0.861±0.16<sub>1</sub> | 0.003±0 | 0±0 | 0.879±0.157 | 25828±237<sub>76</sub> | 2.6±0.5 | 101.8±1.2 |
| | | $q = 3$ | 0±0 | 0.72±0.147 | -0.003±0 | 0±0 | 0.746±0.147 | 3193±1124 | 2.2±0.4 | 103.1±0.6 |
| | Bacteria | $q = 0$ | 0.019±0.008 | 7.216±0.03<sub>6</sub> | 0.046±0.01<sub>6</sub> | 0±0 | 7.114±0.057 | 1826±728 | 1535.9±2.9 | 89.1±3.2 |
| | | $q = 1$ | 0.034±0.01 | 2.952±0.1 | 0.055±0.01<sub>1</sub> | 0±0 | 2.843±0.069 | 3889±1133 | 25.7±3.2 | 81.2±3.8 |
| | | $q = 2$ | 0.042±0.022 | 2.192±0.13<sub>2</sub> | 0.069±0.02<sub>5</sub> | 0±0 | 2.055±0.086 | 2975±1052 | 13.5±2.5 | 79.7±7.3 |
| | | $q = 3$ | 0.043±0.025 | 1.945±0.13<sub>9</sub> | 0.071±0.03 | 0±0 | 1.806±0.089 | 3498±1078 | 10.7±2.0 | 80.1±8.1 |
| | Viruses | $q = 0$ | 0.14±0.002 | 4.556±0.04<sub>8</sub> | 0.161±0.00<sub>5</sub> | 0±0 | 4.461±0.063 | 28480±127<sub>58</sub> | 310.1±24.3 | 32.3±2.7 |
| | | $q = 1$ | 0.021±0.008 | 2.898±0.04<sub>9</sub> | 0.015±0.02<sub>3</sub> | 0±0 | 2.886±0.11 | 3803±1713 | 21.8±1.3 | 99.6±10.7 |
| | | $q = 2$ | 0.021±0.021 | 2.024±0.07 | 0.018±0.03 | 0±0 | 1.986±0.124 | 3839±1820 | 9.4±0.5 | 105.0±19.4 |
| | | $q = 3$ | 0.016±0.024 | 1.711±0.08<sub>7</sub> | 0.022±0.02<sub>6</sub> | 0±0 | 1.641±0.102 | 4916±1740 | 6.686±0.3 | 105.0±19.3 |

From Table S7 and Table 2, some very interesting numbers that directly answer the question raised in the article title are worthy of emphasis here. Since no statistically significant differences were detected between tissue types, there was no need to distinguish between PT, SN and B samples to estimate the numbers of microbial species (microbial species richness) or MAD (maximum accrual diversity) in terms of the Hill numbers or species equivalents. Due to the nature of MAD, which is estimated from the saturation point of the DAR-PLEC curve (*i*.*e*., the diversity accumulation in terms of Hill numbers), the title question can be converted into how many species there are in human tissue microbiomes. We can depend on the total tissues (PT, SN, B) samples combined to estimate MAD. As to the sequencing protocols, due to potential differences in processing samples and computational pipelines, we do not combine their samples (results). Instead, we treat their estimation results as the range of MAD estimation.

Figure 2 illustrates the potential microbial diversity (Dmax) of pan-tumor microbiome for each taxon (archaea, bacteria, or viruses) and each tissue type (primary tumor [PT], solid normal [SN], or blood [B]), as well as the ‘Total’ of combined tissue types and taxa, at different diversity orders (q=0, 1, 2, 3), under each sequencing protocol (WGS or RNA-Seq). The pan-tumor microbiome concept means that the DAR model is built across cancer types—the microbiomes of all cancer types are combined as the microbial landscape. Therefore, Fig 2 actually shows the potential diversity at the largest pan-tumor scale of tissue microbiomes across all cancer types, including the ‘Total’ that also combined the tissue types besides cancer types. Given the lack of statistically significant differences between tissue types and the saturation nature of *D*_max_ estimation, Figure 2 also illustrates the potential diversity of human tissue microbiomes (the last bar in purple).

**Fig 2.**
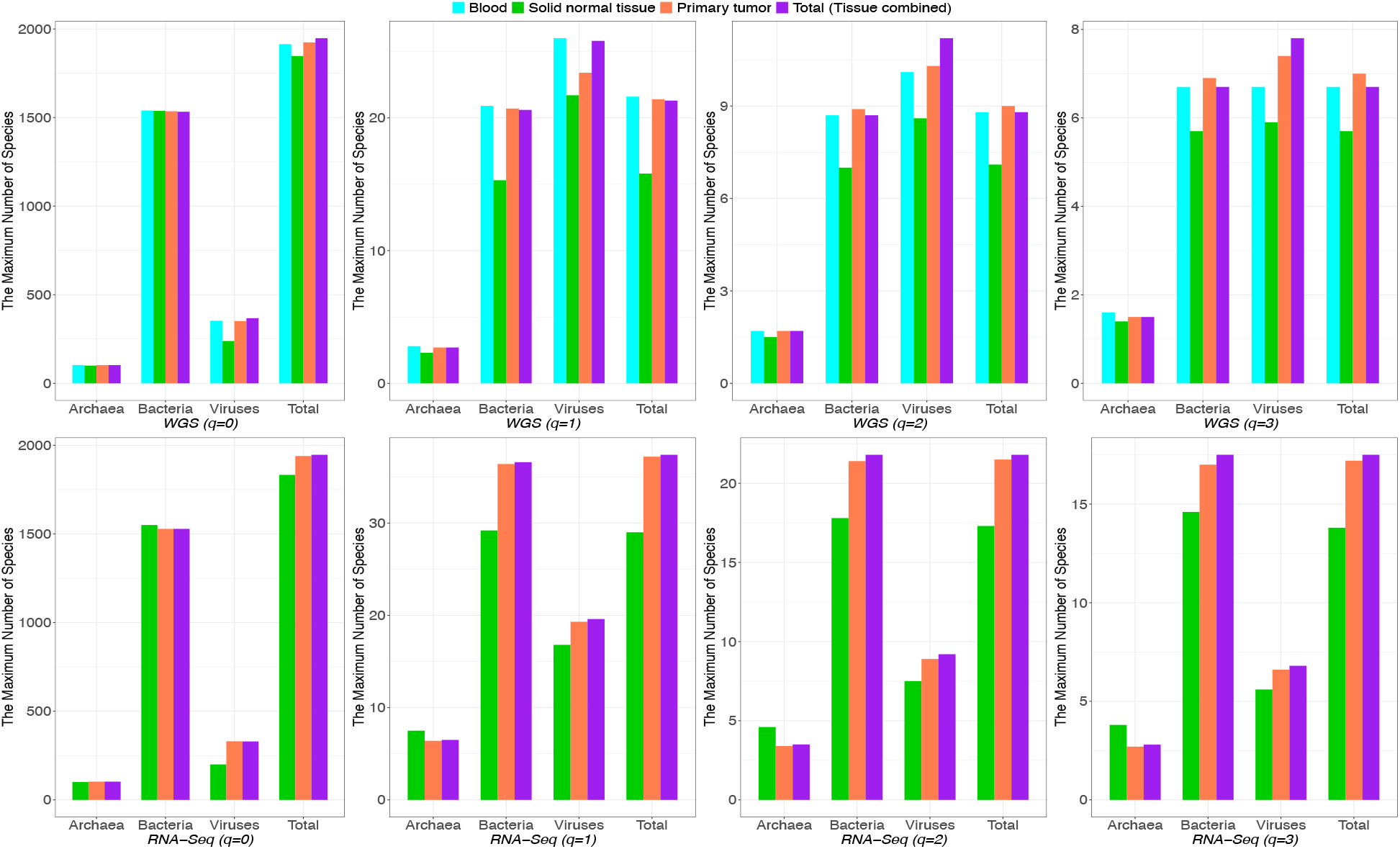
The potential microbial diversity (*D*_*max*_) of pan-tumor microbiome for each taxon (archaea, bacteria, or viruses) and each tissue type (PT, SN, or B), as well as the ‘Total’ of combined tissue types and taxa, at different diversity order (q=0, 1, 2, 3), under each sequencing protocol (WGS or RNA-Seq). The pan-tumor microbiome concept means that the DAR model is built across cancer types—the microbiomes of all cancer types are combined as the microbial landscape and the DAR models were built on the landscape (pan-tumor scale). Given the lack of statistically significant differences between tissue types and the saturation nature of *D*_*max*_ estimation, the last bar (in purple) represents the potential diversity of the human tissue microbiomes.

After the previous rational simplification, from Table S7 we conclude that the species richness (q=0) or the total number of tissue microbial species is between 1946 (estimated from RNA samples) to 1948 (estimated from WGS samples), which are rather close to each other and demonstrate the robustness of the Poore et al. (2020) TCGA datasets used in this study. The categorical breakdown of species richness for taxa are archaea between 102 (WGS) and 103 (RNA), bacteria between 1528 (RNA-Seq) and 1536 (WGS), and viruses between 330 (RNA-Seq) and 368 (WGS). Except for viruses, the numbers from both sequencing protocols are rather close and their differences are negligible.

For diversity order q=1, or the number of species equivalents of typical abundances, the numbers are between 21 (WGS) to 36 (RNA-Seq) or 26 on average across protocols and tissue types. The taxa breakdowns of typical species are archaea between 2.7 (WGS) and 6.5 (RNA-Seq), bacteria 21 (WGS) and 37 (RNA-Seq), and viruses between 20 (RNA-Seq) and 26 (WGS).

For diversity order q=2, or the number of species equivalents of dominant abundances, the numbers are between 21 (WGS) and 22 (RNA-Seq). The taxa breakdowns of dominant species are archaea between 2 (WGS) and 4 (RNA-Seq), bacteria between 9 (WGS) and 22 (RNA-Seq), and viruses between 9 (RNA-Seq) and 11 (WGS).

## Conclusions and Discussion

Regarding the diversity-scaling analysis of the tumor microbiomes, the previous findings can be summarized in the following main conclusions. Except for D_max_, the diversity scaling parameters are generally not significantly different between tissue types and among cancer types (except for q=0 or species richness with RNA-Seq). For D_max_, on average, the differences among tissue types are around 5%, and around 25% (16%-34%) among cancer types. Additionally, the differences in D_max_ among cancer types also vary in terms of sequencing protocol (RNA-Seq < WGS) and tissue types (PT > SN).

This level of diversity analysis performed for tumor microbiomes summarized here mirrors the population-level diversity-disease relationship (DDR) previously reported by Li and Ma (2021), in which they found, based on the analysis of 23 microbiome-associated diseases, that the population-level DDR or p-DDR was only significant in approximately 5% of cases for the parameter D_max_, and was insignificant for other DAR parameters. A significant relationship means that the parameters are different between the disease and health states, or between PT and SN or between PT and B. On this point, both the diversity-cancer relationship (DCR) and general DDR at the host population level show the same pattern—virtually all major DAR scaling parameters are invariant (especially z) except for D_max_, which is variable in approximately 5% of cases. Note that the p-DDR or p-DCR are different from DDR/DCR at the individual host level, or microbial alpha-diversity level, where Ma *et al*. (2019) found that the DDR relationships were significant in approximately 1/3 of the cases they studied. Both studies analyzed the same datasets of 23 microbiome-associated diseases but generated different results. Li and Ma (2021) postulated that it should be the mutual cancellations of individual-level DDR differences (ups and downs) that generated the ‘flat’ or insignificant scaling parameters at the host population level. Here, we believe that the findings on p-DCR in this study simply cast supportive evidence on that previous hypothesis—that mutual cancellations of the ups and downs at the individual host level differences are responsible for the general lack of differences at the host population level.

In this study, our p-DDR level analysis goes beyond single disease or single cancer type; instead, our diversity-scaling analyses were performed at the individual cancer type level (for comparing different tissue types such as PT vs. SN or B) and at the pan-tumor level (across cancer types for comparing different cancer types such as lung vs. breast cancer), respectively. This pan-disease analysis was not performed in previous studies by Li & Ma (2021) and Ma *et al*. (2019) because those microbiome datasets were not from single tissue types—instead they included rather heterogeneous samples from gut, oral, skin, and vaginal microbiomes. In this study, all microbiomes were from human tissues or blood. Although we consider the tissue microbiome samples to be more homogeneous than those analyzed in the previous studies (Ma *et al*. 2019, Li & Ma 2021), there should still be a certain level of heterogeneity among different tissue types. We postulate that the relatively larger D_max_ differences (around 25%) at the pan-tumor level among cancer types, compared to approximately 5% difference at the tumor level, should be attributed to the heterogeneity among different cancer types including different tissues such as lung versus breast tissues.

Given the general lack of differences in major scaling parameters, especially z, at the pan-tumor level analysis, we further built pan-tumor DAR models (Table S7) by combining all disease types. Further tests (Table S8) of the pan-tumor population DAR models (pp-DAR models) revealed no significant differences between tissue types (PT, SN, or B), which prompted us to further build pp-DAR models by combining tissue types (Tables S7 & S8). In Table S7 and Table 2 (summary version), we show that the total species richness, typical species equivalents, and dominant species equivalents of human tissues including blood are approximately 1948, 36, and 22, respectively.

Among the total numbers, the proportions of archaea, bacteria, and viruses are about 5%, 78%, and 17%, respectively, in terms of RNA-Seq. The proportions are slightly higher (about 1%) for bacteria and viruses in terms of the WGS protocol, and that for archaea would be approximately 3% only.

To the best of our knowledge, there is no existing study that has estimated the total (gamma) microbial diversity at the host population level for tumor tissue microbiomes, or actually for any human tissue microbiomes. Since no significant differences were detected between tumor tissues and solid normal tissues (or even blood-derived normal), our results with combined cancer types and tissue types can be considered an estimate of gamma diversity of the human tissue microbiome. Although no similar estimates exist for human tissues, there were similar estimates for non-tissue human microbiomes. In fact, the development of the DAR modeling method extending the classic SAR (species-area relationship) was demonstrated with Human Microbiome Project (HMP) datasets (HMP Consortium 2012; Ma 2018a, 2018b, 2019). We choose two human microbiome sites, skin and gut (stool), to compare with the tissue microbiome. For species richness (q=0), the total species in the tissue microbiome is approximately 1⁄4 (25%) of the gut microbiome (7706) and 1/8 (12.5%) of the skin microbiome (16206). For the number of typical species (q=1), the number of typical species equivalents in tissue microbiomes is 3.5% of gut microbiomes (1020), and 6% of skin microbiomes (605). For the number of dominant species (q=2), the number of dominant species equivalents is 8% of the gut microbiome (272), and 40% of the skin microbiome (55). The higher similarity between tissue and skin microbiomes than between tissue and gut microbiome in terms of dominant species equivalents is somewhat puzzling and deserves further investigation, given that one is ‘internal’ and another is ‘external’ and therefore should be dissimilar. In contrast, for the total species number, the tissue microbiome is indeed more similar to the gut microbiome than to the skin microbiome. Note that our comparisons here are purely based on numbers of species, rather than on compositional comparisons. It is likely that the comparisons are not very informative and instead simply offer relative numbers of species equivalents.

## Data Availability

Availability of datasets
Poore, GD, E Kopylova, Q Zhu, R Knight (2020). Microbiome analyses of blood and tissues suggest cancer diagnostic approach. Nature 579, 567-574. https://doi.org/10.1038/s41586-020-2095-1

## Abbreviations

B: (Blood Derived Normal)
DAR-PL: (DAR Power Law)
D_max_: =Maximal Accrual Diversity (MAD)
DCR: (Diversity-Cancer Relatinship)
MAD: (Maximal Accrual Diversity)
PT: (Primary Tumor Tissue)
SN: (Solid Normal Tissue)
DAR: (Diversity-Area Relationship)
DAR-PLEC: (DAR Power Law with Exponential Cutoff)
DDR: (Diversity-Disease Relationship)
LGD: (Local to Global Diversity Ratio)
p-DDR,: p-DCR (popula6on-level DDR/DCR)

